# Mitigating Hallucinations in Large Language Models: A Comparative Study of RAG-enhanced vs. Human-Generated Medical Templates

**DOI:** 10.1101/2024.09.27.24314506

**Authors:** Anson Li, Renee Shrestha, Thinoj Jegatheeswaran, Hannah O. Chan, Colin Hong, Rakesh Joshi

**Author notes:** These authors contributed equally to the study (Anson Li, Renee Shrestha).

## Abstract

The integration of Large Language Models (LLMs) is increasingly recognized for its potential to enhance various aspects of healthcare, including patient care, medical research, and education. The well-known LLM from Open AI: ChatGPT, a user-friendly GPT-4 based chatbot, has become increasingly popular. However, current limitations to LLMs, such as hallucinations, outdated information, and ethical and legal complications may pose significant risks to patients and contribute to the spread of medical disinformation. This study focuses on the application of Retrieval-Augmented Generation (RAG) to mitigate common limitations of LLMs like ChatGPT and assess its effectiveness in summarizing and organizing medical information. Up-to-date clinical guidelines were utilized as the source of information to create detailed medical templates. These were evaluated against human-generated templates by a panel of physicians, using Likert scales for accuracy and usefulness, and programmatically using BERTScores for textual similarity. The LLM templates scored higher on average for both accuracy and usefulness when compared to human-generated templates. BERTScore analysis further showed high textual similarity between ChatGPT- and Human-generated templates. These results indicate that RAG-enhanced LLM prompting can effectively summarize and organize medical information, demonstrating high potential for use in clinical settings.

## Introduction

Large language models (LLMs), a type of generative artificial intelligence (AI) have the potential to become an effective tool within healthcare. Among LLM-based chatbots, ChatGPT stands out as a prominent example. ChatGPT possesses the ability to “comprehend” and generate human language, which allows it to produce datasets and content derived from existing data on the internet (Birhane et al., 2023; Ray, 2023). The strategic use of prompt engineering techniques and the implementation of Retrieval-Augmented Generation (RAG) when using LLMs can be crucial in delivering responses that are relevant, coherent, and useful to their users.

### What is ChatGPT’s role in medicine?

ChatGPT has the potential to be incorporated into multiple aspects including patient care, medical research, and medical education (Clusmann et al., 2023). Efficient AI systems are paramount to meet the growing demands and challenges in the shortage of the healthcare workforce and the increased administrative burden. However, liabilities in using AI for diagnostic practices and planning treatments have been raised due to accountability and accuracy issues (khan et al., 2023). ChatGPT has shown its extensive medical knowledge capabilities where it has performed well in both medical licensing examinations (such as the USMLE), and in handling medical queries (Gilson et al., 2023; Johnson et al., 2023; Yeo et al., 2023). In combination with its natural language processing capabilities, it has the potential to assist in the communication of medical information with patients by providing simplified summaries within the realm of digital health applications (Dave et al., 2023).

ChatGPT is also used in medical research in all aspects of the research pipeline. It can leverage its extensive medical knowledge to help summarize scientific concepts and existing evidence, accurately and efficiently analyze large datasets, and produce scientific texts that are indistinguishable from human writing (Dave et al., 2023; Ruksakulpiwat et al., 2023; Wong et al., 2023). In its current state, ChatGPT has found its most significant utility in supporting medical education. It can provide summaries, presentations, translations, step-by-step explanations, and guides for a variety of complex medical subjects (Dave et al., 2023).

### Limitations to Generative AI Chatbots like ChatGPT

The main disadvantage of LLMs is the phenomenon of “hallucination”, where it may generate non-existent or false content. This can pose a huge risk to patients seeking medical advice and could contribute to the dissemination of false medical information to the public (Alkaissi & McFarlane, n.d.; Birhane et al., 2023; Liu et al., 2023). ChatGPT is also not currently updated with the latest medical findings and guidelines at a consistent pace, leaving their “extensive knowledge” to be quite static (Gilson et al., 2023; Liu et al., 2023; Tang et al., 2023). Within medical research, ChatGPT exhibits shortcomings in oversimplifying complex scientific concepts, and failing to capture any false information and contextual cues that are readily discernible to human researchers (Birhane et al., 2023). Although ChatGPT adheres to the EU’s AI ethical guidelines aimed to tackle these ethical and legal complications, it may not be enough to prevent the misuse of ChatGPT in a healthcare setting in its current state (Dave et al., 2023). Using AI in a medical setting can elicit ethical and legal complications such as inaccurate and biased outputs, fabricated content, copyright infringements, lack of human accountability, and possible violations of patient privacy resulting from data collection (Wang et al., 2023).

### What is the significance of Retrieval-Augmented Generation (RAG)?

RAG is a framework that can improve LLM responses by referencing external supplemented sources of information outside of its internal training data sources. Implementing RAG provides the LLM with up-to-date, reliable and specific information while giving users access to its sources to allow for the verification of information. Theoretically, RAG reduces “hallucinations”, a phenomenon where Chat-GPT can produce false sources and content (Chen et al., 2016). Although there have been numerous studies looking into the accuracy and reliability of the medical content produced by ChatGPT, there is still a lack of studies that evaluate the use of RAG and its potential to mitigate the limitations mentioned above.

### Utilization of RAG-integrated LLMs in Generating Medical Templates

This study aims to assess the accuracy and effectiveness of RAG-integrated ChatGPT responses in organizing medical information. The information will be organized in a consultation template. The results from this study can provide evidence for the use of LLMs in helping physicians streamline their note-taking process during patient care, and optimizing time efficiency (Alissa et al., 2022; Becker et al., 2010). Physicians also often rely on structured templates to guide their consultations, especially for complex or uncommon cases. Therefore, the approach used in this study can also present a new method of creating dynamic physician-consultation aids that adapt to rapidly changing medical guidelines. Moreover, it can serve as a valuable education tool for students in healthcare, allowing them to gain familiarity with the processes of patient consultation, diagnosis, and treatment.

## Methods

### Data Source for Template Making

The LLM generative AI system used for this study was a RAG-integrated GPT-4 through the OpenAI interface. The medical information was structured in an Electronic Medical Record (EMR) template format. The latest practical guidelines taken from the British Medical Journal (BMJ) were uploaded as PDFs as the source for the ChatGPT model to retrieve information.

A comprehensive array of 15 BMJ practical guidelines was employed to investigate diseases and conditions within the specialized domains of ophthalmology, cardiology, and dermatology. The ophthalmology disease guides include acute conjunctivitis, age-related macular degeneration, amblyopia, angle-closure glaucoma, and astigmatism. The cardiology diseases studied include acute heart failure, chronic atrial fibrillation, diabetic cardiovascular disease, essential hypertension, and unstable angina. Lastly, the dermatology diseases guides include acne vulgaris, basal cell carcinoma, eczema, melanoma, and squamous cell carcinoma. This provided use cases for various diseases, spanning diverse medical specialties, to assess the applicability of employing ChatGPT for templating practical guidelines about commonly encountered medical conditions.

### ChatGPT Prompting

To prompt ChatGPT to create templates, few-shot prompting, directional stimulus prompting, and self-refine prompting methods were applied. Few-shot prompting entailed providing ChatGPT with detailed instructions and illustrative examples to guide its output generation process. This ensures enough context and unambiguity in the outputs generated. Directional stimulus prompting involved providing keywords and phrases to guide the LLM toward the desired output. Self-refining prompting included critiquing the outputs generated by ChatGPT to improve the outputs provided (Figure 1).

**Figure 1.**
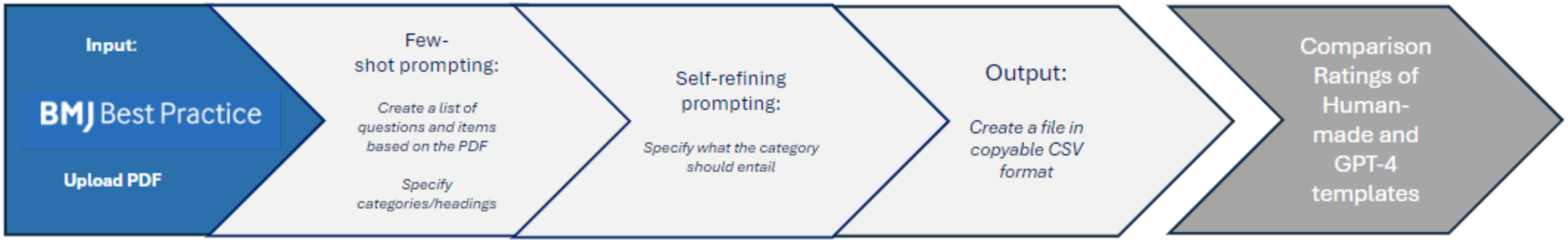
Overview of using RAG and Evaluation of ChatGPT-4 Templates. ChatGPT-4 is prompted to read the BMJ PDF file and generate a list of questions based on what a physician would consider important. This output is then refined through self-refining prompting of categories for the final output. The final output is then prompted to be a copyable CSV file. This file is then evaluated against a Human-generated template of the same condition.

The first prompt is a thorough detailed request that includes all the instructions necessary to create the templates as copyable CSV outputs. This includes a request to read through and understand the uploaded disease guidelines and provide a list of questions and statements regarding the disease under the categories of epidemiology, etiology, pathophysiology, history of patient illness, observations/examinations, classification, grade (severity), investigations, differential diagnosis, and treatments. To get the most detailed response, an exact number of questions/statements required under each category was also included in the prompt, as this will vary in the default LLM output, depending on the complexity of the disease. The model was also given an example of patient-physician interaction during a doctor’s visit, along with the questions commonly asked for diagnosis and treatment planning. This is intended to ensure that the questions and statements in the template closely resemble those encountered during a consultation. After providing the specific requests for each category, the final request is for ChatGPT to generate the output in a CSV format that can easily be exported as an Excel sheet file.

### Mitigating ChatGPT’s Limitations through Self-Refining Prompting

Due to the complexity and length of the source file and requested prompt, ChatGPT usually only provides a limited number of categories (2-3) at once. Self-refining prompting was used to request ChatGPT to continue producing the remaining categories and to potentially fix any errors in its previously generated responses. Reiteration of category-specific phrases and instructions is crucial to reinforce ChatGPT’s understanding of the precise expectations for its output generation. Upon completion of the generation of outputs for each distinct category by the LLM, a final self-refinement prompt is utilized to combine these outputs into a comprehensive CSV format. It is imperative to specify in this last step that the contents previously generated should remain unaltered, ensuring the preservation of detailed information and mitigating the risk of any reduction in detail due to the complexity of the request (Figure 2).

**Figure 2:**
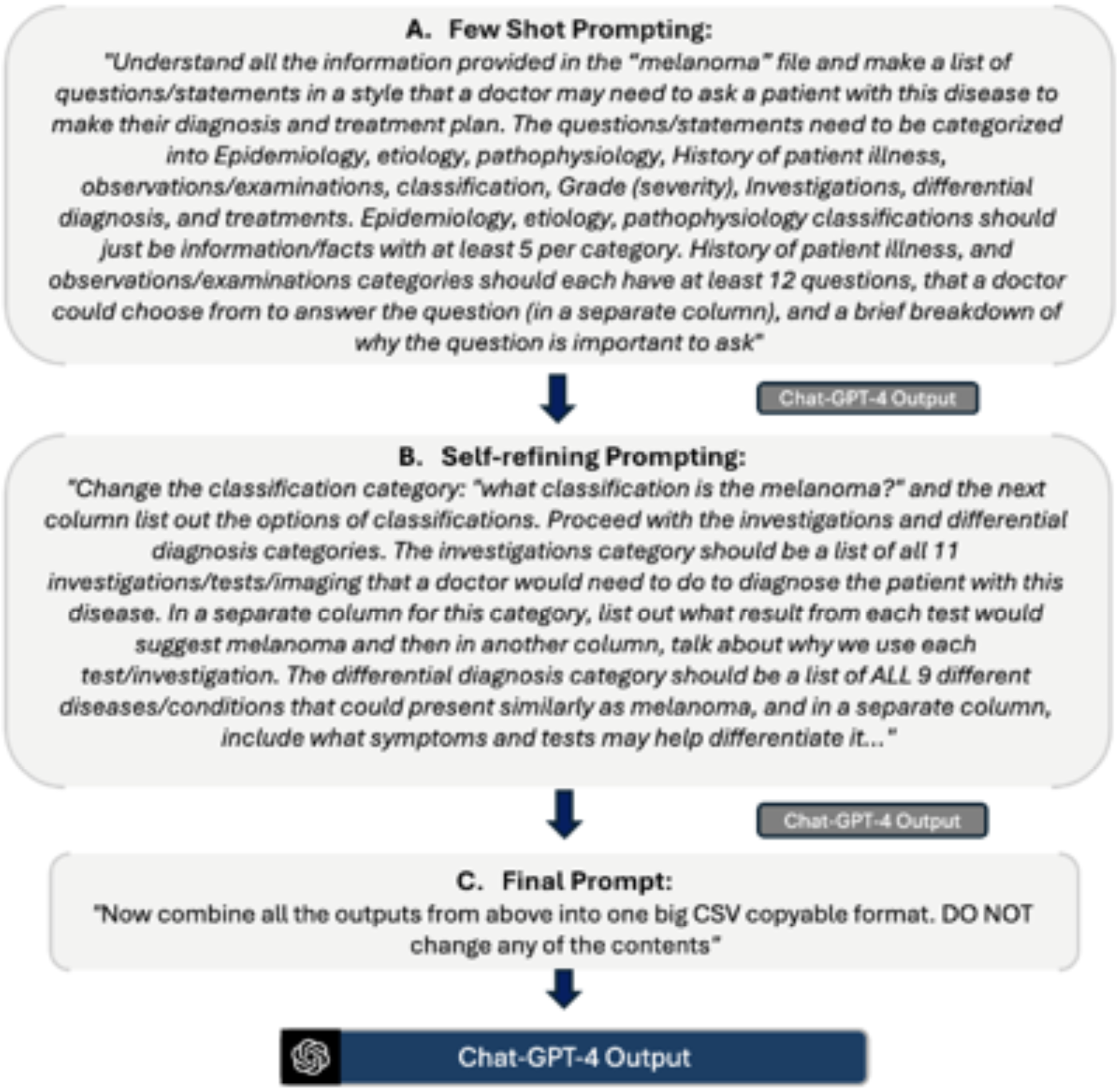
Flow chart containing experts initial prompts and output taken from melanoma file. ChatGPT-4 is then able to refer to the previous output to modulate the next output after receiving input on modifications to make.

### Human Generated EMR Templates

As shown in Table 1, for each ChatGPT-generated EMR template, an analogous EMR template was also created manually from the same BMJ PDF by a non-physician and approved by a physician. This was used to serve as the reference standard against which the ChatGPT-generated templates were evaluated (Table 2).

**Table 1:** Rules for Each Category in Templates.

| Headers | Rules and Regulations Prompted |
| --- | --- |
| Epidemiology, Etiology, Pathophysiology | - Provide facts that were useful for a physician to know for their consult under each respective category. |
| History of Patient Illness, Observations/Examinations | - Generate a set of questions and examinations deemed essential for a thorough consultation. answer options, and brief explanations of the importance of each question or examination were requested in separate columns<br>- Example of how a typical question under this category was also provided to ChatGPT to ensure questions were generated from a third-person point-of-view.<br>- Ordering of the questions and examinations listed was also specified to mirror the order in which a physician would typically ask. For example, start with pressing concerns and symptoms and end with detailed historical and familial background information. |
| Classifications, Grade (Severity) | -<br>- Provide the classification groups, the types of classification forth corresponding groups classification options and information of those classification types in separate columns<br>- similar process was conducted with the grade (severity) category, where a similar format was requested, but for information pertaining to the severity of the disease. |
| Investigation | -<br>- Specifically requested the tests and imaging needed to be conducted to diagnose the patients.<br>- For each test or imaging modality, corresponding results indicative of the disease were detailed, along with a comprehensive explanation justifying the necessity of conducting each test or imaging procedure.<br>- These components were also requested to be arranged into distinct columns. |
| Differential Diagnosis | -<br>- A comprehensive list of alternative diseases sharing symptomatic similarities with the target disease was requested to be compiled. For each listed condition, distinguishing symptoms and diagnostic tests were requested to be arranged into separate columns.<br>- Emphasis was placed on utilizing only the diseases and conditions provided in the uploaded source file to ensure adherence to specified parameters and avoid reliance on external knowledge. |
| Treatment | - Requested to list all the treatments used, name, dose, regimen, and route of administration (oral, topical, intravenous/subcutaneous, or other).<br>- Information on when, and why a certain treatment should be used is also requested in separate columns.<br>- Specify that conservative treatments (lifestyle, diet, exercise, etc.) should also be listed to reduce the emphasis that ChatGPT has on medical and surgical interventions. |

**Table 2:** Average Ratings of Usefulness and Accuracy by Type of Disease.

|  | Usefulness | Accuracy |
| --- | --- | --- |
| Cardiology Disease |  |  |
| ChatGPT Templates | 3.80 | 4.60 |
| Human Templates | 3.53 | 4.53 |
| Ophthalmology Diseases |  |  |
| ChatGPT Templates | 4.40 | 4.87 |
| Human Templates | 4.00 | 4.47 |
| Dermatology Diseases |  |  |
| ChatGPT Templates | 4.20 | 4.40 |
| Human Templates | 3.60 | 4.40 |

**Table 3:** Cohen’s Kappa Scores for Usefulness.

| Cohen's Kappa Scores (ChatGPT) |  |  |  |
| --- | --- | --- | --- |
| Voters | Kappa | Standard Error | 95% CI |
| Voter 1-2 | 0.339 | 0.141 | 0.064 to 0.615 |
| Voter 2-3 | 0 | 0 | 0 |
| Voter 1-3 | 0.102 | 0.074 | -0.043 to 0.248 |
| Cohen's Kappa Scores (Human) |  |  |  |
| Voter 1-2 | -0.179 | 0.151 | -0.475 to 0.115 |
| Voter 2-3 | -0.114 | 0.170 | -0.449 to 0.220 |
| Voter 1-3 | 0.029 | 0.207 | -0.378 to 0.436 |

## Statistical Analysis

### Likert Scale

To evaluate the accuracy of the content generated by ChatGPT in comparison to the gold standard of human generated EMR templates, a panel of three physicians scored both categories of templates. They rated both ChatGPT- and human-generated templates across 15 diseases using a 5-point Likert scale (1—Very inaccurate, 2—Inaccurate, 3—Equally accurate and inaccurate, 4— Accurate, 5—Very accurate). To gauge the usefulness of the templates for the physicians, the physicians also assessed them on a 5-point Likert scale (1—Very useless, 2—Useless, 3—Equally useless and useful, 4—Useful, 5—Very useful). The averages of the Likert scales calculated for the ChatGPT-generated templates were then compared with the averages of the Likert scales calculated for the human-generated EMR templates using the dependent sample t-test.

### BERTScore

BERTScore is a metric used when token-level similarities are investigated. Tokens are individual pieces of text the model processes, which can be whole words or parts of words. BERTScoring was used to programmatically compare human- and ChatGPT-generated templates. This metric offers an alternative to calculating cosine similarity, between embeddings, by focusing on token-level comparison rather than whole-document embeddings. It aligns individual tokens between two texts and uses their cosine similarity to compute a score. This approach can be more reflective of actual textual similarity, especially in contexts like evaluating detailed medical documents where precise language is crucial.

Briefly, paired documents, the LLM- and Human-generated templates for the same disease, were loaded onto a Python (ver 3.7) Colab notebook, whose codes can parse and tokenize the text, use the BERT language model to handle the parsed tokens, and then uses the <BERT-score> function to analyze and compare the two documents at the token level.

## Results

### Human Evaluations

Among the three raters, the mean Usefulness Likert Scale score was 4.13 for the ChatGPT-generated templates compared to 3.71 for the Human-generated templates. In the subset of cardiology disease templates, the ChatGPT-generated templates received a mean score of 3.80, while the human-generated templates received a mean score of 3.53. For ophthalmology disease templates, the ChatGPT-generated templates were rated with a mean score of 4.40, compared to 4.00 for the human-generated templates. Similarly, the dermatological disease templates generated by ChatGPT scored a mean of 4.20, whereas the human-generated templates scored 3.60.

For accuracy, the mean Accuracy Likert Scale score was 4.62 for the ChatGPT-generated templates and 4.47 for the Human-generated templates. Breaking each disease category down, the cardiology disease generated by ChatGPT received a mean score of 4.60 while the Human-generated templates received a score of 4.53. For the ophthalmology disease templates, the ChatGPT-generated templates were rated with a mean score of 4.87 while the Human-generated templates received a mean score of 4.47. In the dermatological disease category, both ChatGPT-generated and human-generated templates achieved an identical mean score of 4.40.

Overall, human evaluations indicated that ChatGPT-generated templates consistently received higher mean scores for usefulness and accuracy across various disease categories than human-generated templates (Table 4; Figure 3).

**Figure 3:**
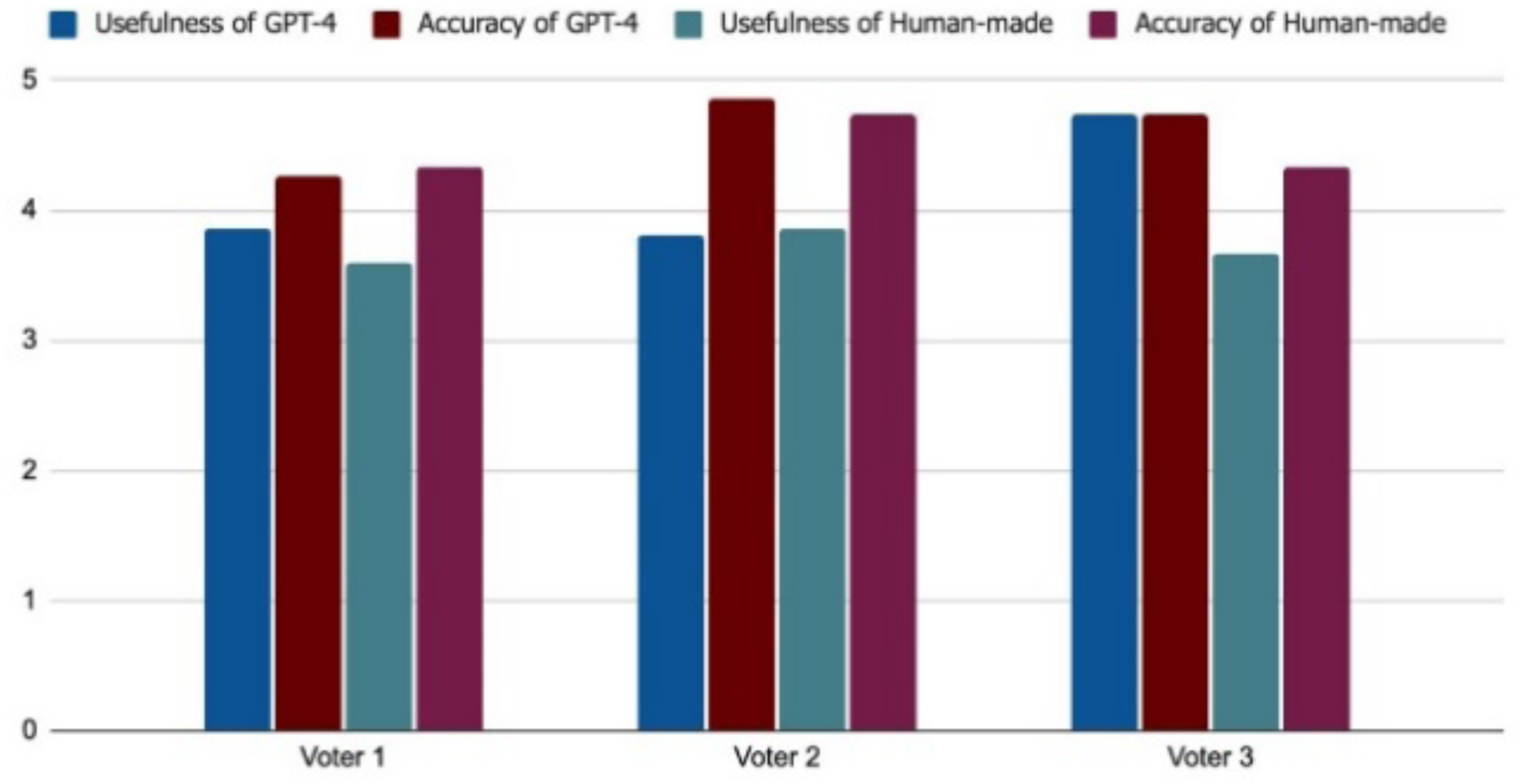
Human physician voter ratings (n = 3) on the usefulness and accuracy of RAG-LLM-Generated and Human-Generated templates.

**Table 4:** Cohen’s Kappa Scores for Accuracy.

| Cohen's Kappa Scores (ChatGPT) |  |  |  |
| --- | --- | --- | --- |
| Voters | Kappa | Standard Error | 95% CI |
| Voter 1-2 | 0.129 | 0.099 | -0.064 to 0.322 |
| Voter 2-3 | -0.154 | 0.082 | 0.313 to 0.006 |
| Voter 1-3 | -0.012 | 0.127 | -0.260 to 0.236 |
| Cohen's Kappa Scores (Human) |  |  |  |
| Voter 1-2 | -0.304 | 0.199 | -0.695 to 0.086 |
| Voter 2-3 | 0.178 | 0.179 | -0.171 to 0.528 |
| Voter 1-3 | -0.382 | 0.152 | -0.679 to -0.084 |

### Interrater Analysis

Cohen’s kappa was employed to evaluate inter-rater reliability. A score is assigned based on the level of agreement between two raters, where a score of 1 indicates perfect agreement and a score of 0 indicates agreement no better than chance (McHugh, 2012). For the Usefulness Likert Scale scores for ChatGPT-generated templates, Raters 1 and 2 had a kappa value of 0.34 (SD=0.14), Raters 2 and 3 had a kappa value of 0 (SD=0), and Raters 1 and 3 had a kappa value of 0.10 (SD=0.07). For the usefulness scores for Human-generated templates, Raters 1 and 2 had a kappa value of -0.18 (SD=0.15), Raters 2 and 3 had a kappa value of -0.11 (SD=0.17), and Raters 1 and 3 had a kappa value of 0.03 (SD=0.21) (Table 5; Figure 4).

**Figure 4:**
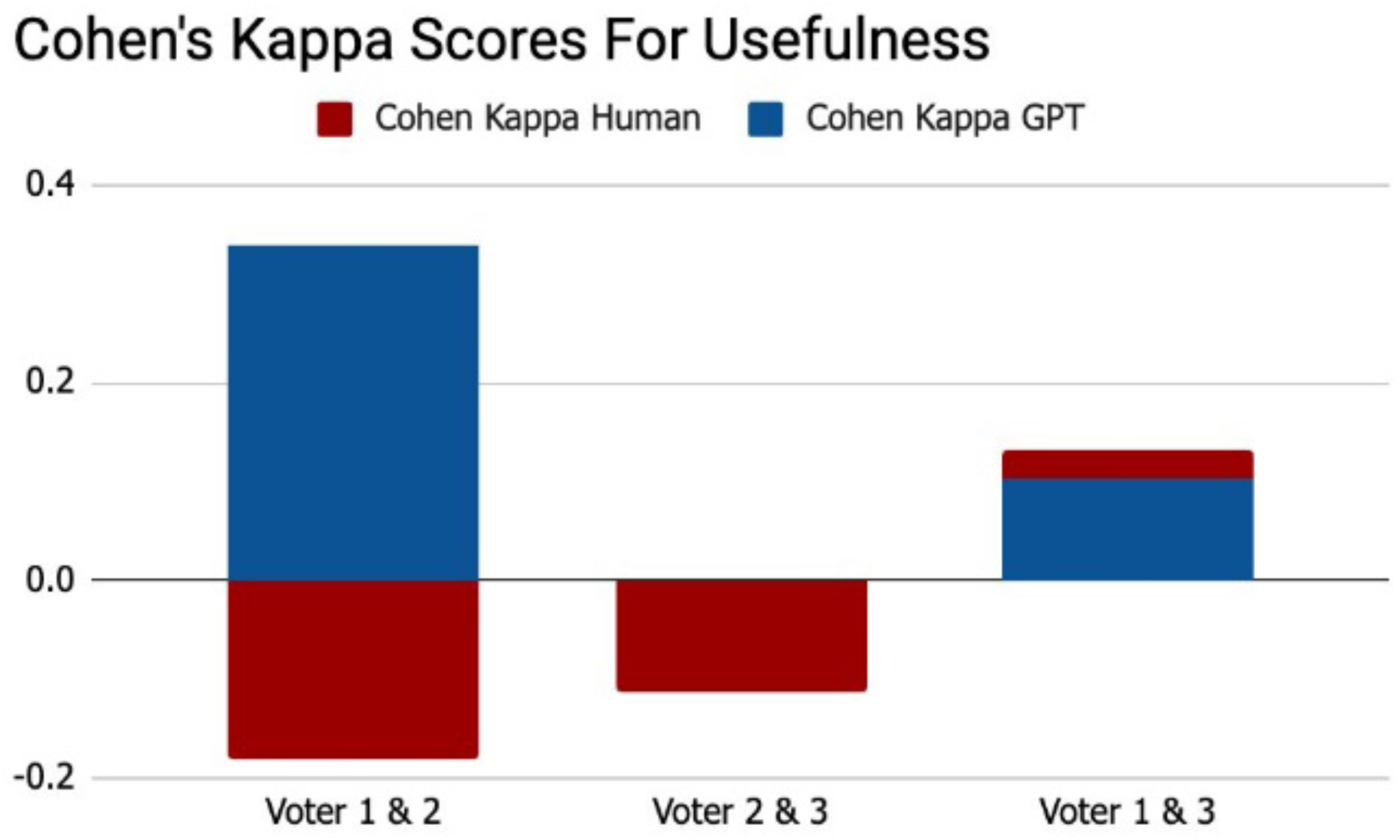
Cohen’s kappa scores. for usefulness, to compare human intra-rater reliability.

**Table 5:** BERTScores for Comparison of ChatGPT- and Human-Generated Templates. For each condition, the BERT Score was calculated. This method evaluates precision, recall, and F1 score, serving as benchmarks for assessing the quality of the generated text. It can be observed that the precision and recall have small ranges.(A) Ophthalmology diseases; (B) Dermatology diseases; (C) Cardiology diseases

| Condition | f1 | Precision | Recall |
| --- | --- | --- | --- |
| A |  |  |  |
| Acute Conjunctivitis | 0.843 | 0.842 | 0.844 |
| Age-related macular degeneration | 0.830 | 0.819 | 0.841 |
| Amblyopia | 0.850 | 0.852 | 0.849 |
| Angle-closure glaucoma | 0.855 | 0.855 | 0.856 |
| Astigmatism | 0.834 | 0.828 | 0.839 |
| Average | 0.842 | 0.839 | 0.846 |
| SD | 0.012 | 0.016 | 0.007 |
| B |  |  |  |
| Acne Vulgaris | 0.834 | 0.829 | 0.839 |
| BCC | 0.837 | 0.835 | 0.839 |
| Eczema | 0.831 | 0.828 | 0.834 |
| Melanoma | 0.852 | 0.850 | 0.854 |
| SCC | 0.853 | 0.857 | 0.849 |
| Average | 0.842 | 0.840 | 0.844 |
| SD | 0.010 | 0.013 | 0.008 |
| C |  |  |  |
| Acute Heart Failure | 0.821 | 0.817 | 0.824 |
| Chronic Atrial Fibrillation | 0.826 | 0.828 | 0.825 |
| Diabetic Cardiovascular Disease | 0.851 | 0.852 | 0.851 |
| Essential Hypertension | 0.833 | 0.841 | 0.825 |
| Unstable Angina | 0.844 | 0.852 | 0.838 |
| Average | 0.835 | 0.838 | 0.833 |
| SD | 0.013 | 0.015 | 0.012 |

For the Accuracy Likert Scale scores for the ChatGPT-generated templates, Raters 1 and 2 had a kappa value of 0.13 (SD=0.10), Raters 2 and 3 had a kappa value of -0.15 (SD=0.08), and Raters 1 and 3 had a kappa value of -0.01 (SD=0.13). For the usefulness scores of Human-generated templates, Raters 1 and 2 had a kappa value of -0.30 (SD=0.20), Raters 2 and 3 had a kappa value of 0.18 (SD=0.18), and Raters 1 and 3 had a kappa value of -0.38 (SD=0.15). These results demonstrate variability in inter-rater reliability, with generally low kappa values across both ChatGPT-generated and human-generated templates (Table 6; Figure 5).

**Figure 5:**
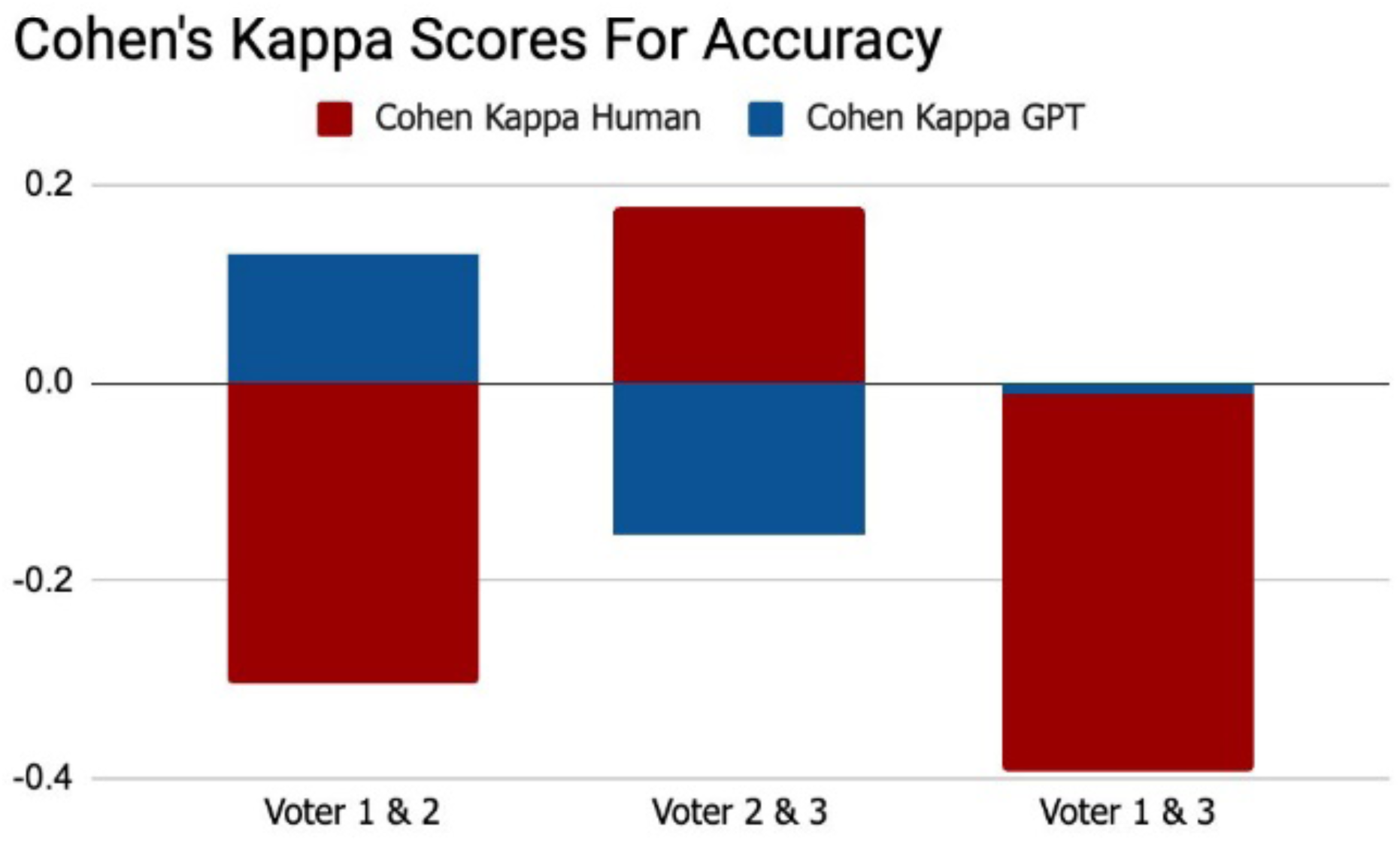
Cohen’s kappa scores, for accuracy, to compare human intra-rater reliability.

The BERTScores were averaged across all disease template pairs to provide a comprehensive similarity measure between human-generated and ChatGPT-generated templates. The BERTScore evaluation encompassed three key metrics: precision, recall, and F1 score. Precision evaluates the ChatGPT-generated templates’ ability to exclude irrelevant information compared to the human-generated templates, while recall assesses their effectiveness in retaining relevant content. Both precision and recall scores were consistently high, with precision values between 0.8171 to 0.8519, and recall values between 0.8241 to 0.8489.

The F1 scores then combine precision and recall to measure how well the ChatGPT-generated templates capture relevant information from the human-generated templates. Across the different medical conditions, the F1 scores ranged from 0.8206 to 0.8504, indicating a high degree of overall similarity between the human- and ChatGPT-generated templates, with minor variations across different medical conditions. These BERTScore metrics indicate that the ChatGPT-generated templates closely and objectively mirrored the content of the human-generated templates, effectively incorporating a substantial amount of relevant information while introducing minimal irrelevant content.

Specifically, the templates for ophthalmological and dermatological diseases exhibited high similarity and consistency in their average F1 scores, with values of 0.8424 (SD =0.0108) and 0.8416 (SD = 0.0103), respectively. In contrast, templates for cardiological diseases were slightly less similar and displayed greater variability, with an average F1 score of 0.8352 (SD = 0.0128). Dermatology disease templates achieved the highest average precision score of 0.8400 (SD = 0.0131), whereas cardiology disease templates had the lowest average precision score of 0.8379 (SD = 0.0152). Furthermore, ophthalmology disease templates recorded the highest average recall score of 0.8459 (SD = 0.0068), while cardiology disease templates had the lowest average recall score of 0.8326 (SD = 0.0120).

The statistical analysis indicated that ChatGPT-generated templates closely mirrored the content of Human-generated templates. This demonstrated a high degree of similarity and effectiveness in incorporating relevant information with minimal irrelevant content.

## Discussions

The results of our study suggest that employing RAG when using ChatGPT holds significant promise for aiding physicians in clinical settings. Through both subjective evaluations by physician raters and objective statistical analyses, ChatGPT demonstrated its ability to accurately summarize and organize medical information based on current clinical guidelines. Out of 45 accuracy ratings, the raters had rated 30 (66.7%) of all the ChatGPT-generated templates as “very accurate” (5 – Likert Scale). This is further supported by statistical evaluation that measured at a BERTScore value of 0.84 in a range of 0 (no semantic similarity) and 1 (perfect semantic similarity), when compared directly to the Human-generated templates. Surprisingly, the average rating given to the ChatGPT-generated templates (4.62) was slightly higher than the rating given to the Human-generated templates (4.47). Additionally, only two Chat-GPT templates were given the lowest rating of “equally accurate and inaccurate” (0.04%), suggesting that ChatGPT, when using RAG, rarely produces incorrect information. This helps address a critical concern regarding the potential risks of ChatGPT in medical settings, as inaccurate information could lead to significant harm.

The raters were also confident in the usefulness of the ChatGPT-generated templates within a clinical setting. Of the 45 ChatGPT-generated templates, 15 were rated as “very useful” (33.3%), whereas nine templates were rated with the lowest score being “equally useful and useless” (2.0%). The median usefulness score was 4. This indicates that most of the templates generated by ChatGPT are well-organized and could significantly benefit physicians. Notably, ChatGPT templates were rated more useful than human-generated templates, with average scores of 4.13 and 3.71, respectively. The rating process was single-blinded which contributes to the conclusion that the ChatGPT-generated templates were comparatively more useful and accurate. Moreover, this indicates the strength of ChatGPT, which is the ability to summarize and organize dense and detailed information that previously only experts would be able to fully decipher. ChatGPT curates the most critical information as prompted by the user, therefore this guided approach would lend itself to more useful and critical information needed by physicians. However, the human-generated templates contained an overwhelming amount of information, indicating that the information from the BMJ PDF was not summarized or organized as efficiently as ChatGPT. Moreover, qualitatively, ChatGPT-generated templates were more uniform.

Despite the encouraging results, there are areas for improvement in ChatGPT’s functionality and output. ChatGPT-4 currently has a limit of 40 messages every 3 hours, which can constrain the productivity of template generation. However, this limitation is expected to be addressed in future updates. In terms of accuracy, although RAG significantly reduces hallucinations, as demonstrated by the precision score values, occasional inaccuracies persist. For instance, ChatGPT sometimes includes diagnostic tests not listed in clinical guidelines, potentially misallocating medical resources and posing risks to patients (Koch et al., 2018). Thus, ChatGPT cannot work independently to curate the templates and would require human input. This may reduce physicians’ confidence in using ChatGPT as an assistive tool. Inaccuracies often require self-refining prompts to ensure adherence to guidelines, requiring the need for human oversight. Therefore, the efficiency of the creation process is hindered. Additionally, the consistency of the CSV file format generated by ChatGPT needs improvement. Variations in column content and number between templates can negatively affect standardization and usability. However, ChatGPT’s potential lies in its capacity for refinement through physician feedback, allowing for better organization of information to meet clinical needs. This capacity is validated by the accuracy and usefulness of RAG-prompted LLMs showing great potential in supporting physicians in their clinical encounters.

Several limitations of this study should be acknowledged. Despite the high scores given by all the human raters for usefulness and accuracy, the small sample size limits the ability to draw definitive conclusions about the overall usefulness and accuracy of the ChatGPT-generated templates. This limitation is further underscored by the variability in agreement among the raters. Critically, the templates that are being evaluated by the three physician raters are from three different medical specialties of which the physicians are not experts. This can minimize the impact of their ratings of usefulness and accuracy as their medical knowledge and practical knowledge of the diseases are limited. There is also a selection bias regarding the physicians recruited to rate the templates. The medical specialties of the disease templates (ophthalmology, cardiology, dermatology) are not fully representative of the nuances, challenges, and specificities required to diagnose and treat diseases of other specialties. Further, to reduce the fatigue effects of the raters, the modest number of disease templates evaluated contributes to this lack of representativeness. Moreover, the results from the version of the chatbot used (ChatGPT-4) cannot be generalized to further versions or of other LLMs which can differ in memory capacity and capabilities. Future research should involve a larger sample size of physician raters with diverse medical backgrounds, use of different LLMs, and a more diverse array of disease templates to better represent the wide variety medical specialties. This will validate the generalizability of these findings across different medical fields.

Another potential limitation is the lack of evaluation of each template’s accuracy, particularly regarding the prevalence of hallucinations. Although the BERTScores did establish a high degree of similarity between the human- and ChatGPT-generated templates, and hallucinations were mitigated by directional prompting, the total amount of prompting and revisions were not considered during the evaluation. This limits the ability to make a definitive conclusion as to how efficient and useful this method is for physicians who have limited time.

The result of this study also has implications for the potential of ChatGPT-4 generated templates to be used for education purposes for medical students. These needs of medical students could potentially be met by the efficiency of ChatGPT-generated templates which are suggested by this study’s results to be accurate and useful. Moreover, in further research medical students should be included in the evaluation process to assess how well these templates facilitate efficiency in the studying process while maintaining academic integrity, potentially enhancing medical education. This dual approach of broadening the clinical scope and exploring educational applications would provide a more comprehensive understanding of the utility and adaptability of ChatGPT-generated templates in various medical contexts. While ChatGPT has shown to be capable of creating templates of diseases of varying complexities across different specialties with directional prompting, further investigation should delve into the accuracy of these templates and evaluation of the frequency of hallucinations.

## Conclusion

The LLM-based chatbot Chat-GPT-4 exhibits strong potential for use within healthcare, hinging on its efficiency in the assessment and summarization of information with customizable prompts, when using Retrieval-Augmented Generation (RAG). The present study investigated the usefulness and accuracy of Chat-GPT-4’s summarization of disease information found in PDFs in ophthalmology, dermatology and cardiology and organization through human prompting. The results show that in comparison to human-made summarized templates, Chat-GPT-4’s template was rated higher for usefulness and accuracy. Also, BERTScore metrics were used to assess the similarity of the ChatGPT-4 template against the human-made templates. This addressed an often- major limitation of the use of ChatGPT which is ‘hallucinations’, in which the chatbot will fabricate information. The BERTScore established that they had a high degree of similarity, indicating that there was little confabulation. However, a key factor to consider when assessing an LLM’s strengths, is its ability to work without human input. However, major ethical and practical issues make directional prompting of LLMs necessary for the desirable output. Therefore, in its current state of advancement, independent analysis and summarization by LLMs for useful medical information is not possible. Nevertheless, AI has great potential for the efficiency of physicians and medical students with its quick turn-around and accuracy in making medical document drafts. As ChatGPT continues to be updated to newer versions with better capabilities, it will become a useful tool to integrate into the medical education and healthcare sectors if practical and ethical issues are addressed.

## Data Availability

All data produced in the present study are available upon reasonable request to the authors

https://www.bmj.com/

## References

1. Alissa, R., Hipp, J. A., & Webb, K. (2022). Saving Time for Patient Care by Optimizing Physician Note Templates: A Pilot Study. Frontiers in Digital Health, 3, 772356. 10.3389/fdgth.2021.772356

2. Alkaissi, H., & McFarlane, S. I. (n.d.). Artificial Hallucinations in ChatGPT: Implications in Scientific Writing. Cureus, 15(2), e35179. 10.7759/cureus.35179

3. Becker, G., Kempf, D. E., Xander, C. J., Momm, F., Olschewski, M., & Blum, H. E. (2010). Four minutes for a patient, twenty seconds for a relative—An observational study at a university hospital. BMC Health Services Research, 10, 94. 10.1186/1472-6963-10-94

4. Birhane, A., Kasirzadeh, A., Leslie, D., & Wachter, S. (2023). Science in the age of large language models. Nature Reviews Physics, 5(5), 277–280. 10.1038/s42254-023-00581-4

5. Chen, C. A., Lin, L., Tsibris, H. C., Li, W.-Q., Li, T. Y., & Qureshi, A. A. (2016). Pilot testing and validation of an atopic dermatitis screening and evaluation questionnaire. International Journal of Dermatology, 55(7), e399–403. 10.1111/ijd.13145

6. Clusmann, J., Kolbinger, F. R., Muti, H. S., Carrero, Z. I., Eckardt, J.-N., Laleh, N. G., Löffler, C. M. L., Schwarzkopf, S.-C., Unger, M., Veldhuizen, G. P., Wagner, S. J., & Kather, J. N. (2023). The future landscape of large language models in medicine. Communications Medicine, 3(1), 1–8. 10.1038/s43856-023-00370-1

7. Dave, T., Athaluri, S. A., & Singh, S. (2023). ChatGPT in medicine: An overview of its applications, advantages, limitations, future prospects, and ethical considerations. Frontiers in Artificial Intelligence, 6, 1169595. 10.3389/frai.2023.1169595

8. Gilson, A., Safranek, C. W., Huang, T., Socrates, V., Chi, L., Taylor, R. A., & Chartash, D. (2023). How Does ChatGPT Perform on the United States Medical Licensing Examination (USMLE)? The Implications of Large Language Models for Medical Education and Knowledge Assessment. JMIR Medical Education, 9, e45312. 10.2196/45312

9. Johnson, D., Goodman, R., Patrinely, J., Stone, C., Zimmerman, E., Donald, R., Chang, S., Berkowitz, S., Finn, A., Jahangir, E., Scoville, E., Reese, T., Friedman, D., Bastarache, J., van der Heijden, Y., Wright, J., Carter, N., Alexander, M., Choe, J., … Wheless, L. (2023). Assessing the Accuracy and Reliability of AI-Generated Medical Responses: An Evaluation of the Chat-GPT Model. Research Square, rs.3.rs-2566942. 10.21203/rs.3.rs-2566942/v1

10. khan, B., Fatima, H., Qureshi, A., Kumar, S., Hanan, A., Hussain, J., & Abdullah, S. (2023). Drawbacks of Artificial Intelligence and Their Potential Solutions in the Healthcare Sector. *Biomedical Materials & Devices (New York*, N.y.), 1–8. 10.1007/s44174-023-00063-2

11. Liu, J., Wang, C., & Liu, S. (2023). Utility of ChatGPT in Clinical Practice. Journal of Medical Internet Research, 25, e48568. 10.2196/48568

12. McHugh, M. L. (2012). Interrater reliability: The kappa statistic. Biochemia Medica, 22(3), 276– 282.

13. Ray, P. P. (2023). ChatGPT: A comprehensive review on background, applications, key challenges, bias, ethics, limitations and future scope. Internet of Things and Cyber-Physical Systems, 3, 121–154. 10.1016/j.iotcps.2023.04.003

14. Ruksakulpiwat, S., Kumar, A., & Ajibade, A. (2023). Using ChatGPT in Medical Research: Current Status and Future Directions. Journal of Multidisciplinary Healthcare, 16, 1513– 1520. 10.2147/JMDH.S413470

15. Tang, L., Sun, Z., Idnay, B., Nestor, J. G., Soroush, A., Elias, P. A., Xu, Z., Ding, Y., Durrett, G., Rousseau, J. F., Weng, C., & Peng, Y. (2023). Evaluating large language models on medical evidence summarization. NPJ Digital Medicine, 6, 158. 10.1038/s41746-023-00896-7

16. Wang, C., Liu, S., Yang, H., Guo, J., Wu, Y., & Liu, J. (2023). Ethical Considerations of Using ChatGPT in Health Care. Journal of Medical Internet Research, 25, e48009. 10.2196/48009

17. Wong, R. S.-Y., Ming, L. C., & Raja Ali, R. A. (2023). The Intersection of ChatGPT, Clinical Medicine, and Medical Education. JMIR Medical Education, 9, e47274. 10.2196/47274

18. Yeo, Y. H., Samaan, J. S., Ng, W. H., Ting, P.-S., Trivedi, H., Vipani, A., Ayoub, W., Yang, J. D., Liran, O., Spiegel, B., & Kuo, A. (2023). Assessing the performance of ChatGPT in answering questions regarding cirrhosis and hepatocellular carcinoma. Clinical and Molecular Hepatology, 29(3), 721–732. 10.3350/cmh.2023.0089

